# Community pharmacists Awareness and Attitude toward Counterfeit medicine in Khartoum Locality: Cross Sectional Survey

**DOI:** 10.1101/2020.10.26.20219501

**Authors:** Wala W. Wagiealla, Shaza W. Shantier, Imad O. Abureid, Elrasheed A. Gadkariem

**Affiliations:** Department of Pharmacy Practice, Faculty of Pharmacy, University of Khartoum, Sudan; Department of Pharmaceutical Chemistry, Faculty of Pharmacy, University of Khartoum, Sudan

**Keywords:** Counterfeit medicines, Pharmacists, Awareness, Attitude, Sudan

## Abstract

**Background:** Counterfeit medicines (CFMs) are a global problem with significant and well-documented consequences for global health and patient safety. Pharmacists, as healthcare professionals, have a vital role in combating CFMs via ensuring the effectiveness and safety of any imported and dispensed medicines.

**Objectives:** The aim of the present study was to assess the extent, awareness and attitude of pharmacists in Sudan towards CFMs.

**Methods:** A cross-sectional study was conducted applying pretested and structured questionnaire and the awareness and attitude were assessed statistically.

**Results:** A total of 229 participants have enrolled in the study. The majority of the respondents (76%) were found to be aware about the term CFMs during practice. 59% reported their ability to distinguish CFMs from the packaging feature and cost. Unavailability of medicine and inadequate regulatory bodies control were mentioned as the main leading factors for the spread of CFMs. 86% of the respondents reported knowing that there are pharmacists who deal with counterfeit medicines believing that they are unethical (76%). 69% of the respondents mentioned pharmacists to have vital role in combating CFMs spread through increasing knowledge and education. 62% of the participants were found to have a fair awareness about CFMs. Furthermore, 56% of them showed a good attitude toward CFMs.

**Conclusion:** Current literature includes gaps in knowledge and attitude towards CFMs. Therefore attention and concentrated efforts are required on the part of the government, drug manufacturers and health care providers’ especially pharmaceutical analysts to ensure that only drugs of acceptable quality reach the patient.

## Introduction

Counterfeit medicines (CFMs) are of lower qualities than their originals, fraudulently manufactured with fake packaging and usually with no or a wrong active pharmaceutical ingredient [1]. Although reporting of counterfeit drugs within WHO is only 15%, CFMs are available all over the world and are most prevalent in developing countries due to weak medicine regulation, control and enforcement [2, 3].

Literature review was conducted to determine the prevalence of CFMs in the United States of America (USA), Europe, Africa and Asia with a focus on the developing countries [4]. The research showed CFM are available worldwide and are most prevalent in developing countries due to weak medicine regulation, control and enforcement. A study was carried out in Saudi Arabia indicated that 34.4% of drugs were counterfeits in 2005 and increase in 2006 to 49.3%. In Africa and Asia the figure exceeds 50% trading in counterfeit and/or substandard drugs annual earnings reached over 35 billion dollars [4, 5].

Sudan is a vast African country surrounded by a number of other African countries (e.g Kynea, Uganda…etc). Most of these African countries suffer from the circulation of such drugs. To combat this problem, these countries have put strategies to control this problem through enforcing strong close and carry out continuous Post Marketing Surveillance (PMS) studies.

The role of pharmaceutical quality control (QC) and Good Manufacturing Practices (GMP) is to assess the quality of active pharmaceutical ingredients (APIs) and excipients [6]. Despite the fact that National Medicine Regulatory Authorities (NMRAs) have been executed in all developing countries, most of them are not completely operational, while the rest at various degrees of foundation.

Many challenges facing authorities, health care providers and National Quality Control Laboratories (NQCL) regarding counterfeit and substandard drugs these include that they are difficult to trace, their spreading can’t be controlled or stopped, their detection, quantification and control need well equipped labs and well-trained trustful personnel and some expired legitimate drugs can be remarked with false new expiry date. In addition to that counterfeiters aim to avoid raising suspicion about the origin and the quality of their products also they take measures that make them slip past the authorities control and ultimately deceive consumers.

Providing enhanced pharmaceutical services would not be enough when the safety and efficacy of medicine may be compromised by the availability of CFM [7]. Therefore, pharmacists’ role in combating CFMs will be via ensuring the effectiveness and safety of any dispensed medicines as per the WHO good pharmacy practice guidelines [8-10].

CFMs are thus raising the challenge against pharmacists’ awareness and professionalism. Various studies have been reported in this concern revealing different levels of awareness about the problem, lack of regulations with the suggestion to design and implement educational programmes for pharmacists [11-13]. Additionally, the lack of knowledge, standards, regulations and laws is affecting the practice and endangering public health [14, 15].

Despite all of that, gaps still exist in current literature regarding pharmacist’s knowledge about counterfeit medications, strategies used by pharmacists to handle counterfeit medications delivered to their practice setting, and how patients are educated about counterfeit medications. Therefore, the aim of this study is to reveal the awareness and attitude of the community pharmacists towards counterfeit medicines in Khartoum, Sudan, their role in educating patients about CFMs and to suggest means in combating this problem.

## Methods

### Setting and study population

The study was conducted in Khartoum locality, Sudan between December 2019 and April 2020. The inclusion criteria were pharmacist working in community pharmacies in Khartoum Locality willing to participate in the study. Closed pharmacies are excluded from the study.

### Sample size and sampling technique

Number of community pharmacies in Khartoum locality was obtained from the Pharmacy Department, Federal Ministry of Heath, Khartoum, Sudan. The sample size was calculated using the following formula [16]:

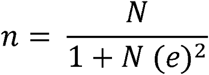

Where: n: sample size e: precision level (0.05); N: population size (954 pharmacies).

Systematic random sampling technique was used to select pharmacies in Khartoum locality,then the pharmacist who was working at the time of data collection was included in the study after assigning the informed consent.

### Questionnaire design and validation

It consisted of demographic characteristics of participants, awareness of CFMs, attitude towards CFMs and recommendations on measures to be taken to control the illicit trade in counterfeit medicines (Supplementary file 1).

The questionnaire was piloted on twenty of the community pharmacists for content validation. It was then simplified, shortened to consume 12-15 minutes. Finally, the questionnaire was assessed through experts in the field of Pharmacy before data collection. Awareness questions were multiple choices and Yes/No questions, while attitude questions were on Likert scale (Agree /Neutral/ Disagree).

### Data collection

Self-administered questionnaire was given to pharmacist working during data collection at the selected pharmacy.

### Data analysis

Data were entered and analyzed using Excel 2016. Descriptive analysis was performed for the questionnaire where frequencies, percentages and graphical representation were reported for all categorical variables. Content analysis was used to analyze the open-ended responses. This involved reading the questions, identifying recurring ideas and categorizing all responses to similar ideas or answers to allow frequency calculation.

Scoring system for awareness was as follows; 5 questions were selected; the correct answer given1 and incorrect answer given 0. A total of 6 points for the awareness classified as (with 0, 1 poor, 2, 3 Fair and 4,5 Good awareness).

Scoring system for attitude was as follows; 10 statements; The positive attitude given 2, Neutral given1 and negative attitude given 0. A total of 20 points(0-7 Poor, 8-13 Fair,14-20 Good).

### Ethical requirements

The ethical clearance (FPEC-03-2019) was obtained from the Ethical Committee, Faculty of Pharmacy, University of Khartoum. The dates and times when the questionnaires were administered were all documented.

All of respondents signed a written informed consent and were guaranteed privacy and confidentiality.

## Results

### Pharmacists’ awareness and attitude

#### Demographic data

The study sample included 229 pharmacists. 138 (60 %) of them were females, the mean age and years of experience are 30.4 and 6.2 years, respectively. Obtained data are summarized in Table 1.

**Table 1.** Socio-demographic Characteristics of the participants (n= 229)

| Demographic data | N | % |
| --- | --- | --- |
| **Gender** |  |  |
| Female | 138 | 60 |
| Male | 91 | 40 |
| **Qualifications** |  |  |
| Bachelor | 186 | 81 |
| Master | 43 | 19 |
| **Years of Experience** |  |  |
| <5 | 114 | 49.8 |
| 5-10 | 77 | 33.6 |
| >10 | 38 | 16.6 |

### Awareness about CFMs

One hundred seventy-three (76%) of participants define that counterfeit medicines are of unknown source, 101 (44%) of bad quality, 84 (37%) with no FDA approval and 61 (27%) as ones containing wrong ingredients. Asking the respondents from which source they became aware of CFMs, 164 (72%) answered during practice, 57 (25%) in university, only 5 (2%) from campaigns and 2 (1%) conference. About CFMs availability in Sudan and areas expected to be found, 200 (87%) of the respondents know that CFMs are available in urban and rural areas in Sudan, while 29 (13%) are not.

The majority of respondents 198 (86 %) reported knowing that there are pharmacists who deal with counterfeit medicines and they are aware of that. The respondents believed that various actions should be taken against those pharmacists (Figure 1).

**Figure 1:**
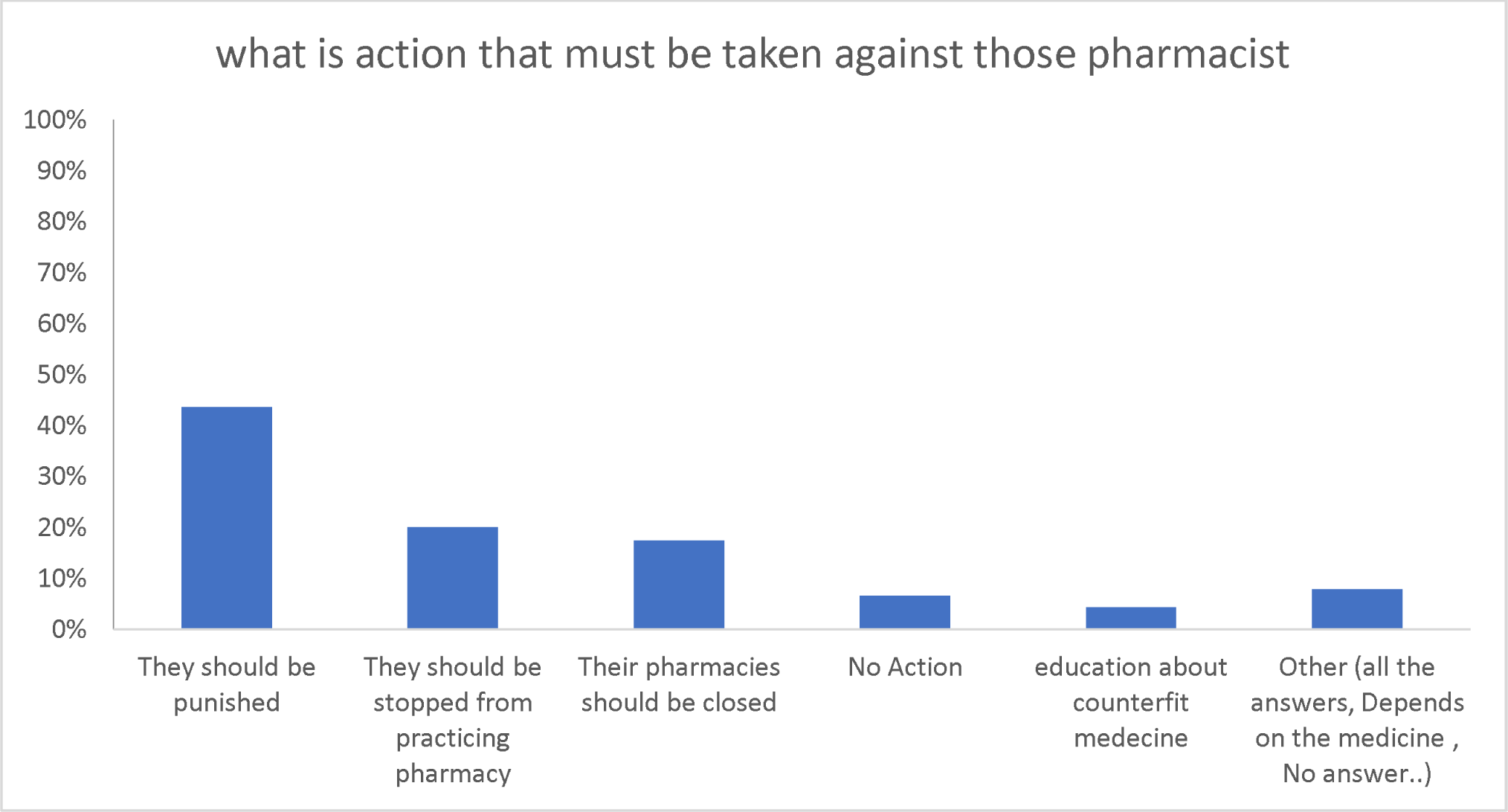
Actions against pharmacists dealing in CFMs.

The respondents reported different characters to distinguish between the CFMs and the originals (Table 2). They also reported various factors leading to the spread of CFMs (Figure 2). In regards to the source of CFMs, 166 (72%) of the respondents reported Egypt as the main origin of CFMs, 55 (24%) mentioned India and 8 (4%) answered China.

**Figure 2:**
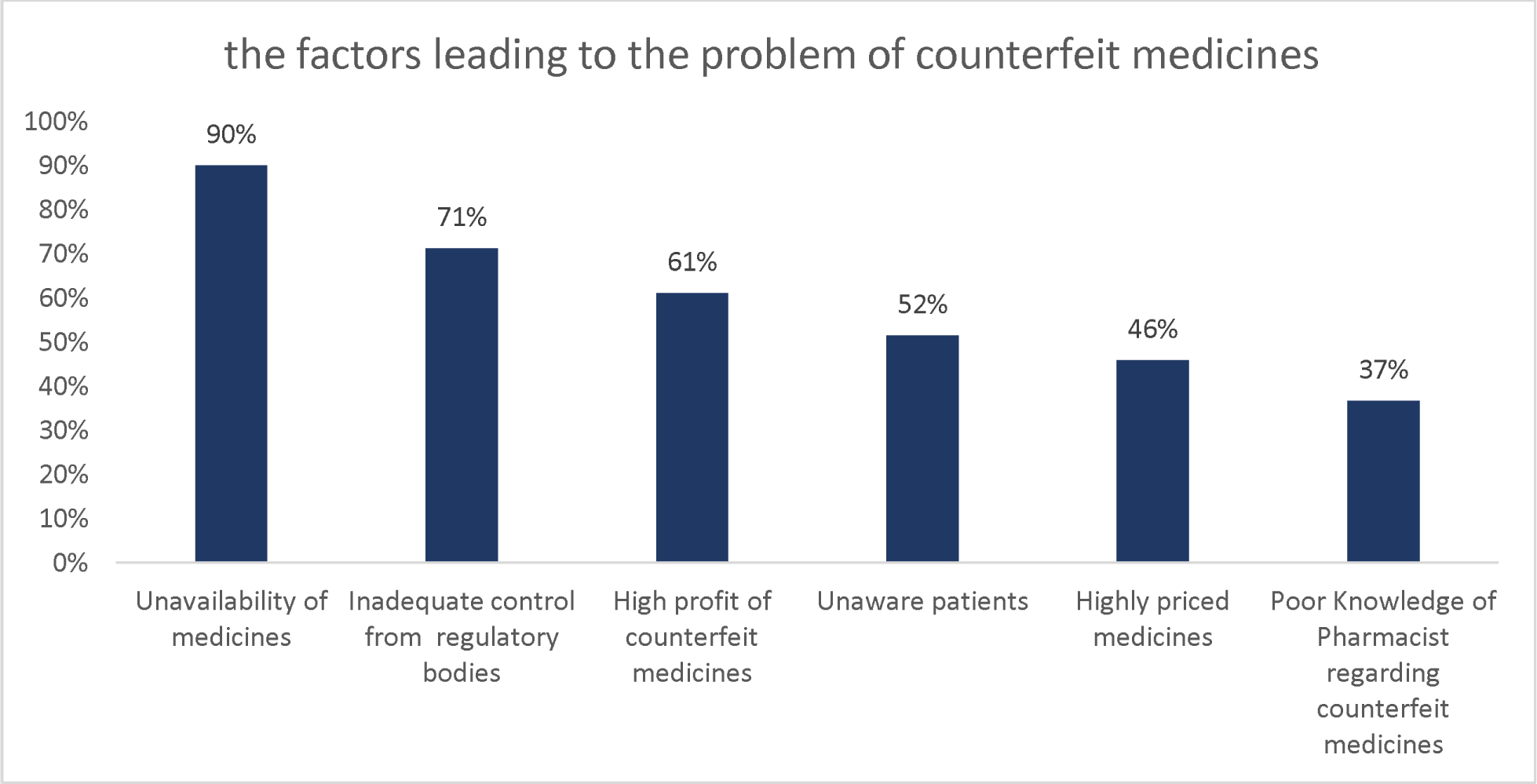
Factors leading to CFMs spread.

Using the scoring system, the obtained results indicated a fair level of awareness in 62% of the respondents, while the good and poor levels of awareness were represented by 10% and 28% of the respondents, respectively.

### Pharmacist’s attitude towards CFMs

Fifty-three (23%) of the respondents reported receiving complains about CFMs. Table 3 summarizes the actions that had been taken towards these complains.

**Table 2.** Characteristics to distinguish between CFMs and Originals.

| Character | N | %* |
| --- | --- | --- |
| Cost (N=229) | 136 | 59 |
| Feature of packaging (N=229) | 133 | 58 |
| Feature of suppliers (N=229) | 96 | 42 |
| Effect of medicine (N=229) | 73 | 32 |
| Package insert information (N=229) | 61 | 27 |

\* Total is more than 100% as more than one answer was possible

**Table 3.** Respondents’ action towards patients’ complains.

|  | Yes |  | No |  |
| --- | --- | --- | --- | --- |
|  | N | % | N | % |
| Informed the company | 5 | 9% | 48 | 91% |
| Informed regulatory bodies | 3 | 6% | 50 | 94% |
| Told the patient to use it | 4 | 8% | 49 | 92% |
| Changed it with another brand name | 18 | 34% | 35 | 66% |
| Asked the patient about the source from which it was bought | 29 | 55% | 24 | 45% |
| You did not do any action | 5 | 9% | 48 | 91% |

Different classes of medicines were reported to be of high risk for counterfieting. 114 (50%) described antimalrials as of high risk, 96 (42%) antihypertensives, 80 (35%) antibiotics, 73 (32%) antidiabetes, 110 (48%) narcotics, 32 (14%) OTC and 42 (18%) GIT medicines. Unavailability and high demand were among the main reasons behind mentioning these drugs as being at high risk of counterfeiting.

The respondents also reported that pharmacists would play a potential role in combating the problem through different measures as shown in Figure 3.

**Figure 3:**
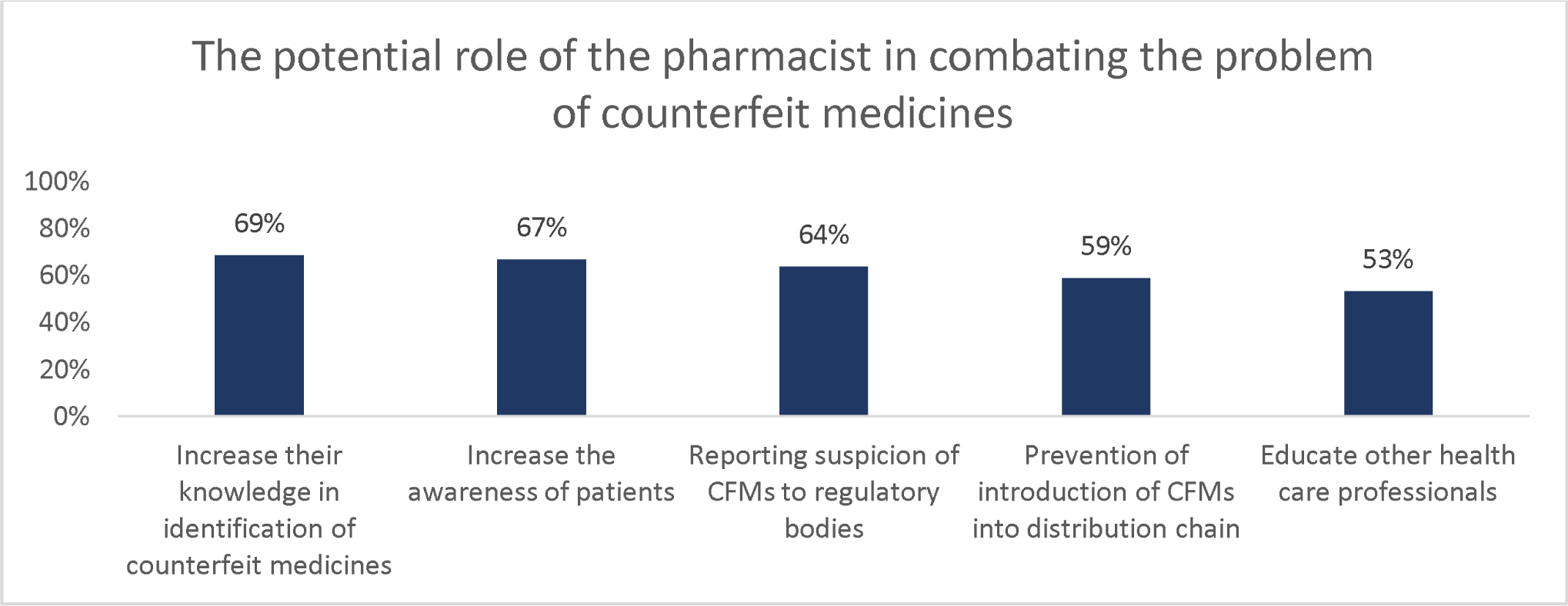
Potential roles to combat CFMs problem.

Pharmacists’ attitude was further assessed using statements describing the quality, risk, experiences, and CFMs’ price. Obtained data are shown in Figure 4 as agree, neutral and disagree based on the respondents’ view.

**Figure 4:**
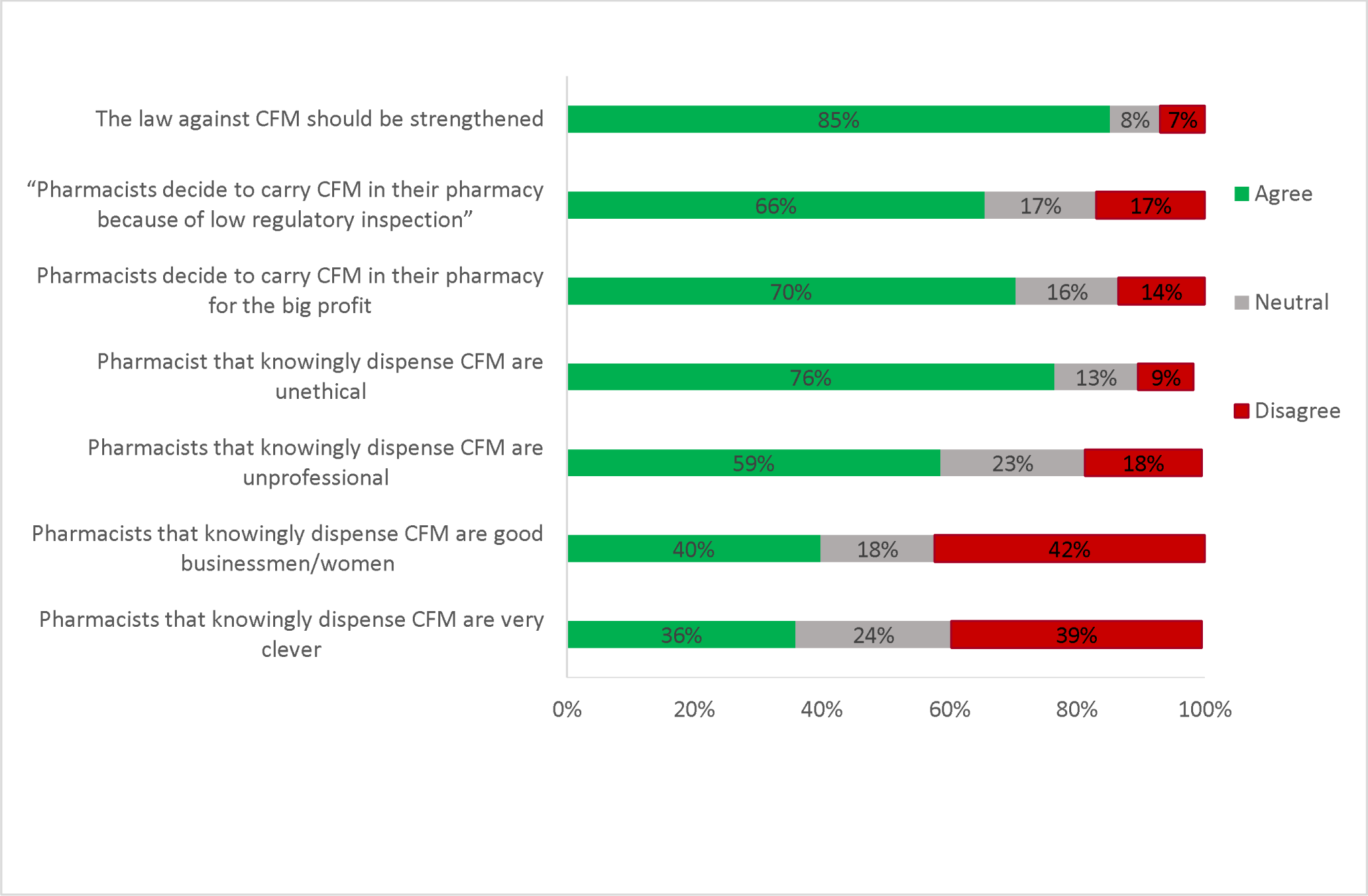
Respondents’ views and experience towards CFMs (N=229)

Regarding the overall attitude, a good attitude was found in 56% of the respondents, while poor and fair attitude were shown in 6% and 38% of the respondents, respectively. No significant correlation was found between awareness, attitude and the tested variables utilizing different statistical tests (Fisher exact test, Kruskal Wallis nonparametric test and Chi square test).

## Discussion

The main public health and economic consequences of the growing trade in counterfeit medicines is affecting poor people in developing countries with limited regulatory and enforcement capacity much more than in the developed countries, with their regulated and relatively transparent supply chains. Clearly, there is work to be done from a structural perspective as the exact extent of this problem, and therefore how best to combat it, is unknown. The WHO is working with the African Union to improve health coverage across Africa, and FDA has also developed a reporting system for suspicion about CFMs through med watch [4, 17].

In this study, the pharmacist’s awareness and attitude have been explored in Sudan and assessed in order to come up with substantial measures and recommendation to help combating the widely spread CFMs issue.

The overall of awareness and attitude of participants was on fair level in comparison to a study conducted in Iran, which reported that participant’s Knowledge was low [18].

In our study, no significant association was found between attitude and any demographic characteristics while in Iranian study there was a significant relation of attitude with age and gender with increasing age and females having high attitude. With regard to awareness no significant association was found between awareness and participants demographic which is the same reported by Iranian pharmacists.

Most of participants defined CFMs are of unknown source followed by, of bad quality which is also reported by a study conducted in Lebanon to assess awareness and views of pharmacists toward CFMs (n=223) [19]. FDA defined CFMs as fake medicine which might be contaminated or contain the wrong or no active ingredient [17], this was only answered by 27% of participants in this study indicating that the precise definition is not known by most pharmacists in Khartoum locality.

High percentage of participants reported that they became aware about the term CFMs from practice and one quarter during study in university indicating that CFMs are not included in the curriculum of most faculties of Pharmacy in Sudan; the same observed by Sholy [19]. Thus, as suggested by a study conducted in California [11], academic institute should have a key role in educating healthcare professionals about CFMs.

The high percentage of participants knows that CFMs are available in the market in Sudan in both urban and rural indicating pharmacists are aware of the spread of the problem.

China and India were believed to be the main sources of CFMs, which have been mentioned also by Sholy et al, 2017 as the main source in Lebanon.

The action that must be taken against pharmacists who deal with CFMs ranging from punishment to closeness of their pharmacies was compared to Lebanon study.

WHO stated that CFMs can be identified by packaging, checking the manufacture, expiry dates, and medicine look [20]. This supports the findings of our study as the participants reported that they identify CFMs packaging and features of suppliers.

Antimalarials and Antibiotics were the most mentioned classes of medicines at high risk for counterfeiting which agrees with WHO stating that Anti-malarials and Antibiotics are amongst the most commonly reported falsified medical products. 60% of the anti-infective drugs in Asia and Africa are containing outside their pharmacopeial limits [21]. Also, this study indicated that Narcotics and Antihypertensive are subjected highly to counterfeiting.

Unavailability of medicines, poor regulation and high profit of CFMs are the most reported factors leading to the problem. These findings were supported by reported studies mentioning that lack of effective regulation, weak legislative and enforcement bodies, cost and unavailability of medicines lead to spread of CFMs [22-24].

The study also indicated that pharmacist can play a vital role in combating CFMs through increasing their own knowledge and educating patients. These results contradict California pharmacist perspective which not strongly agrees that level of knowledge as a barrier to combat the problem, patients’ awareness and other health care professional’s knowledge about CFMs [11]. Improving communication between healthcare professionals and authorities, reporting suspicion and prevention of CFMs entry into distribution chain are of greater impact. These are in addition to interventions suggested by Fadlallah, 2016 in order to combat the problem which include regulatory bodies’ measure, drug laws, legislation, public awareness and education.

The limitation of the present study is that the population may not appear diverse enough; as respondents were reported to be mainly from Khartoum city. Additionally, the cross-sectional design of the study not allows generalization of the findings to all community pharmacists in Sudan. Further studies may be needed in other cities and rural areas of Sudan to assess the awareness and attitude towards CFMs. Despite this, and due to the limited information available regarding pharmacist knowledge and experiences with counterfeit medications, our study would be the groundwork for future studies in exploring the possible implications of counterfeit medications and brings attention to this potential problem faced by the pharmacy profession..

## Conclusion

The present study concluded that most of the respondents were aware about CFMs with positive attitude towards it. Practice was reported as the main source of information by the respondents. No significant correlation between variables was reported. In addition, it highlighted the essential role that pharmacists can play in combating the spread of CFMs and ensuring the safety of the patients. However, great efforts are required to continue educational programs and to improve the communication between pharmacists and regulatory bodies in order to provide more resources and dissemination of information about CFMs.

## Data Availability

All dataset supporting this article is available within the article

## Acknowledgements

Authors acknowledge Faculty of Mathematical sciences, University of Khartoum, Sudan for the sample calculation and statistical analysis. Authors are also thankful for the surveyors, Mr. Mohamed A., Ms. Roaa E., Mr. Khalid M., Ms. Sara O., Ms. Rana E., Ms. Ayat I., who helped in the data collection and entry.

## Conflict of interest

Authors declare no conflict of interest

## Data availability

All dataset supporting this article is available within the article and its supplementary files.

## Funding

This research is fully funded by the Ministry of Higher Education and Scientific Research, Sudan

## Author contribution

W.W wrote the paper draft, conducted part of data collection and entry; S.S. wrote the abstract and conclusion, edit and revise the draft, conducted part of data entry; I.A. and E.A. revise the final draft.

## Notes

### Competing Interest Statement

The authors have declared no competing interest.

### Author Declarations

Ethical Committee (FPEC-03-2019), Faculty of Pharmacy, University of Khartoum.

